# Predictors of hospital admission in young patients with COVID-19

**DOI:** 10.1101/2025.05.22.25327949

**Authors:** Reshvinder S. Dhillon, Matthew Weise, Brian H. Wrotniak, Keith Cross

## Abstract

**Background:** The impact of age, race, and comorbidities on COVID-19 severity in younger populations is not well understood. This study aimed to identify predictors of hospital admission in young patients with COVID-19.

**Methods:** We conducted a retrospective analysis of 2658 COVID-19 patients under 36 years old from March 1 to August 6, 2020, using data from HEALTHeLINK, a regional health information system in western New York. Patients were divided into pediatric (0-19 years) and young adult (20-36 years) groups. We evaluated associations between risk factors and hospital admission using recursive partitioning and linear regression.

**Results:** The study included 2131 young adults and 527 pediatric patients. In young adults, race was the strongest predictor of admission, followed by BMI. African Americans with BMI > 23 had the highest admission rate (63%, p<0.001). Asian race predicted outpatient management regardless of BMI. Smoking and hypertension were less significant predictors, while gender, diabetes, respiratory conditions, and sickle cell disease were not significant. In the pediatric population, race was also the primary predictor of admission, with African Americans having higher admission rates than Whites and Asians. BMI percentile was not a predictor in pediatric patients.

**Conclusions:** Race strongly predicted hospital admission in young COVID-19 patients, with African Americans most likely to be admitted and Asians least likely. For African American young adults, BMI > 23 was an additional strong predictor. A simple decision tree incorporating age, race, and BMI can help identify young patients least likely to require inpatient management.

## Background

Severe Acute Respiratory Syndrome Coronavirus 2 (SARS 2/COVID-19), a novel coronavirus which originated in the Wuhan city of China in late 2019, subsequently spread across the globe. The rapidity and extent of the spread lead the World Health Organization (WHO) to release a statement on March 11^th^ 2020, labelling the outbreak a pandemic.^1^

As of July 2021, there were over 191,000,000 cases and 4,102,000 deaths globally.^2^

The clinical signs and symptoms of COVID-19, the disease process caused by SARS-CoV-2, varies with multiple systems involved in both the acute and convalescent phases.^3^ Respiratory symptoms include cough, shortness of breath, dyspnea, acute respiratory distress syndrome, and respiratory failure. However, multiple other systems, including gastrointestinal, hematologic, cardiac, and neurologic, are commonly involved. In addition to the variety of presentations, the severity of certain symptoms and complications, such as those in Multisystem Inflammatory Syndrome in Children or coagulation derangements in adults, contributes to morbidity and mortality.

Many adults with more severe clinical courses had underlying comorbidities, including diabetes, hypertension, and coronary artery disease.^4–6^ These comorbidities are all known complications of obesity. Indeed, severe obesity (defined as BMI ≥ 40) has been identified as an independent risk factor associated with both risk of hospitalization and risk of intensive care unit admission and mechanical ventilation.^7,8^ At the same time, obesity (defined by the WHO as BMI ≥ 30) has been identified as a potential epidemiologic risk factor for increased morbidity due to COVID-19.^7–9^ Indeed, a study of patients hospitalized with COVID-19 in Shenzhen, China showed that obese patients were more likely to develop severe pneumonia and have a more severe clinical course.^10^

Most of these studies and their resulting insights concern older patients; this is understandable given the high prevalence of morbidity and mortality of COVID-19 in this population. As more young patients are now being seen with COVID-19, we sought to evaluate the significance of obesity and other factors as predictors of severe disease specifically in pediatric and young adult patients.

## Methods

### Study Design and Data collection

This was a retrospective, cohort analysis of COVID-19 positive patients in the Western New York region. All patients analyzed were between the ages of 2 and 35. Data for these patients was collected from HEALTHeLINK. The HEALTHeLINK system is a collaboration among hospitals, physicians, health plans and other health care providers in eight counties of western New York State to securely exchange clinical information and consented patient medical records.

Extracted HEALTHeLINK data was de-identified to insure patient privacy. This data included patients that were positive for COVID-19 up to August 8, 2020. Available information included basic demographic data -- age, gender, race, body mass index (BMI), height, smoking status, underlying medical conditions (including asthma, hypertension, sickle cell, and diabetes), length of hospital stay, ventilator requirement and death.

Initially 3177 patient records were obtained. 517 records were excluded from the final analysis due to duplication (same patient acquired multiple COVID tests), or due to incomplete demography such as absent BMI, race or hospital course information.

Patents were separated into 2 cohorts based on age: 2-19 and 20-35 years old. The BMI for the pediatric population was calculated based on percentile using the WHO chart for age and sex while the BMI for the adults was determined using the formula weight (kg) / height (meters^2^) ^16^. For the pediatric patients, BMI percentiles are as follows; underweight was classified as a BMI below 5^th^ percentile, healthy weight 6^th^ – 84^th^ percentile, overweight 85^th^ – 94^th^ percentile and obese greater than 95^th^ percentile.

### Statistical analysis

Descriptive characteristics for participants were computed to characterize the study sample. Categorical variables (e.g., gender, race, ethnicity) were reported as proportions in percentages, and continuous level variables (e.g., age) as means and standard deviations.

The analytic plan for the primary study aim involved a 3-tier approach. First, a single univariate linear (for continuous) or logistic (for categorical) regression model examined the unadjusted association between the outcome measure and obesity. Next, the outcome variable was regressed on obesity after adjustment for potential confounding variables: age, gender, race, smoking status. Finally, underlying medical conditions (including asthma, hypertension, sickle cell, diabetes), were added to the adjusted model to assess their influence as potential mediators of the association between obesity and COVID-19 outcome.

Unadjusted and adjusted odds ratios (OR) and 95% confidence intervals (CI) for outcomes were calculated, and statistical significance was assessed by using a Wald’s test. All statistical tests were 2-tailed and a P value < 0.05 considered statistically significant. Analyses were conducted using SYSTAT version 13 (SYSTAT Software, 2004).

The data were also analyzed using recursive partitioning, using the CHAID growing method. Admission was the dependent variable while race, BMI, age, gender, diabetes, asthma, respiratory disease, hypertension, sickle cell and smoking status were the independent (predictor) variables.

## Results

Data was available for 3316 patients who tested positive for COVID-19 from the start of the pandemic through August 8, 2020. 658 patients were excluded for age less than 2, incomplete information (i.e lacking BMI, race), and duplicate information (same patient acquiring multiple COVID-19 tests). The resulting 2658 were divided into young adults (20-35 years) and pediatric (2-19 years) populations.

### Pediatric and Young adult results

Patient demographics are listed in **Table 1**. 527 pediatric patients and 2131 young adults were analyzed.

**Table1:** The demographic of young adult and pediatric population with COVID-19.

|  | Young Adults | Pediatrics |
| --- | --- | --- |
| Cases Analysed | 2131 | 527 |
| Mean Age | 28.3 ± 4 | 13.1±5 |
| Female | 63% | 50% |
| Male | 37% | 50% |
| Race |  |  |
| White | 50% | 40% |
| African American | 29% | 28% |
| Asian | 9% | 23% |
| Native American/Alaskan | 2% | <1% |
| Smoking status |  |  |
| Any form of smoking | 12% | 4% |
| Never smoke | 60% | 96% |
| Co-morbidities |  |  |
| Diabetes | 4% | 1% |
| Respiratory disease | 8% | 11% |
| Hypertension | 4% | 0.40% |
| Sickle cell | 0.20% | 0.60% |
| Admission status |  |  |
| Inpatient | 60% | 24% |
| Outpatient | 40% | 76% |
| Median Length of stay | 1.4 SD±2 | 1 SD±1.1 |
| BMI |  |  |
| Underweight | 1% | 3% |
| Healthy | 28% | 55% |
| Overweight | 30% | 17% |
| Obese | 40% | 25% |

Inpatient population of young adults and pediatric were analyzed and are shown in **Table 2**. The racial distribution of pediatric inpatient population differed from the general pediatric cohort, with African Americans making up most of the inpatient population (50%) while only being 28% of the pediatric population. The young adult inpatient cohort mirrors the pediatric finding and shows a higher proportion of African Americans being admitted while a lower proportion of Asians were admitted, when compared to the general statistic of this cohort.

**Table 2:** The analysis of inpatient and outpatient population of each population cohort in regard to BMI, Gender, Smoking status, race and co-morbidities.

|  | Young Adult |  | P value |  | Pediatrics |  | P value |
| --- | --- | --- | --- | --- | --- | --- | --- |
|  | Inpatient | Outpatient | Young Adult |  | Inpatient | Outpatient | Pediatrics |
| BMI (N = 1562) |  |  | 0.00 | N = 462 |  |  | 0.091 |
| Underweight | 1% (4) | 2%(16) |  |  | 4% (4) | 3% (10) |  |
| Healthy weight | 22% (149) | 19% (294) |  |  | 45%(51) | 58%(203) |  |
| Overweight | 26% (173) | 41% (275) |  |  | 21%(24) | 15%(53) |  |
| Obese | 51% (339) | 38% (292) |  |  | 31%(35) | 24%(82) |  |
| Gender (N=2122) |  |  | 0.006 | n=525 |  |  | 0.012 |
| Female | 66% (570) | 60% (758) |  |  | 60%(77) | 47%(186) |  |
| Male | 34% (293) | 40% (501) |  |  | 40%(52) | 53%(210) |  |
| Smoking Status (N=1509) |  |  | 0.5 | n=297 |  |  | 0.067 |
| Any form of smoking | 17%(103) | 15%(137) |  |  | 5%(4) | 4%(9) |  |
| Never smoke | 83%(513) | 85%(756) |  |  | 95%(78) | 96%(206) |  |
| Race (N= 1694) |  |  | <0.00001 | n=397 |  |  | 0.00 |
| White | 49%(374) | 59%(557) |  |  | 47%(52) | 41%(118) |  |
| African American | 45%(339) | 21%(196) |  |  | 50%(55) | 23%(65) |  |
| Asian | 3%(19) | 15%(140) |  |  | 3%(3) | 34%(97) |  |
| Native American | 1%(6) | 3%(27) |  |  | 0%(0) | 0%(1) |  |
| Others | 3%(19) | 2%(17) |  |  | 1%(1) | 2%(6) |  |
| Co-morbidities N=304 |  |  | 0.0095 | n=66 |  |  | 0.04 |
| Diabetes | 43 | 46 |  |  | 4 | 1 |  |
| Respiratory disease | 80 | 37 |  |  | 22 | 34 |  |
| Hypertension | 57 | 37 |  |  | 1 | 1 |  |
| Sickle cell | 4 | 0 |  |  | 3 | 0 |  |

### Pediatric population analysis

In a two-way analysis, there was no difference in the distribution of BMI groups between races. When analyzing inpatient and outpatient COVID-19 positive patients, no statistical difference was found between weight classes in the pediatric population. No significant difference was also found when looked at co-morbidities, smoking status, and length of stay between these weight classes. Cardiac complications, such as supraventricular tachycardia, have been documented in pediatric COVID-19 patients and may warrant closer evaluation in future studies.^19^

When analyzed using recursive partitioning (**Figure 1**), we found race to be the greatest factor for determining admission. African Americans had the highest likelihood of admission, while Asians had the lowest. For the African American group, age was found to be the greatest subsequent predictor for admission, while for the White population, BMI was more significant than age.

**Figure 1:**
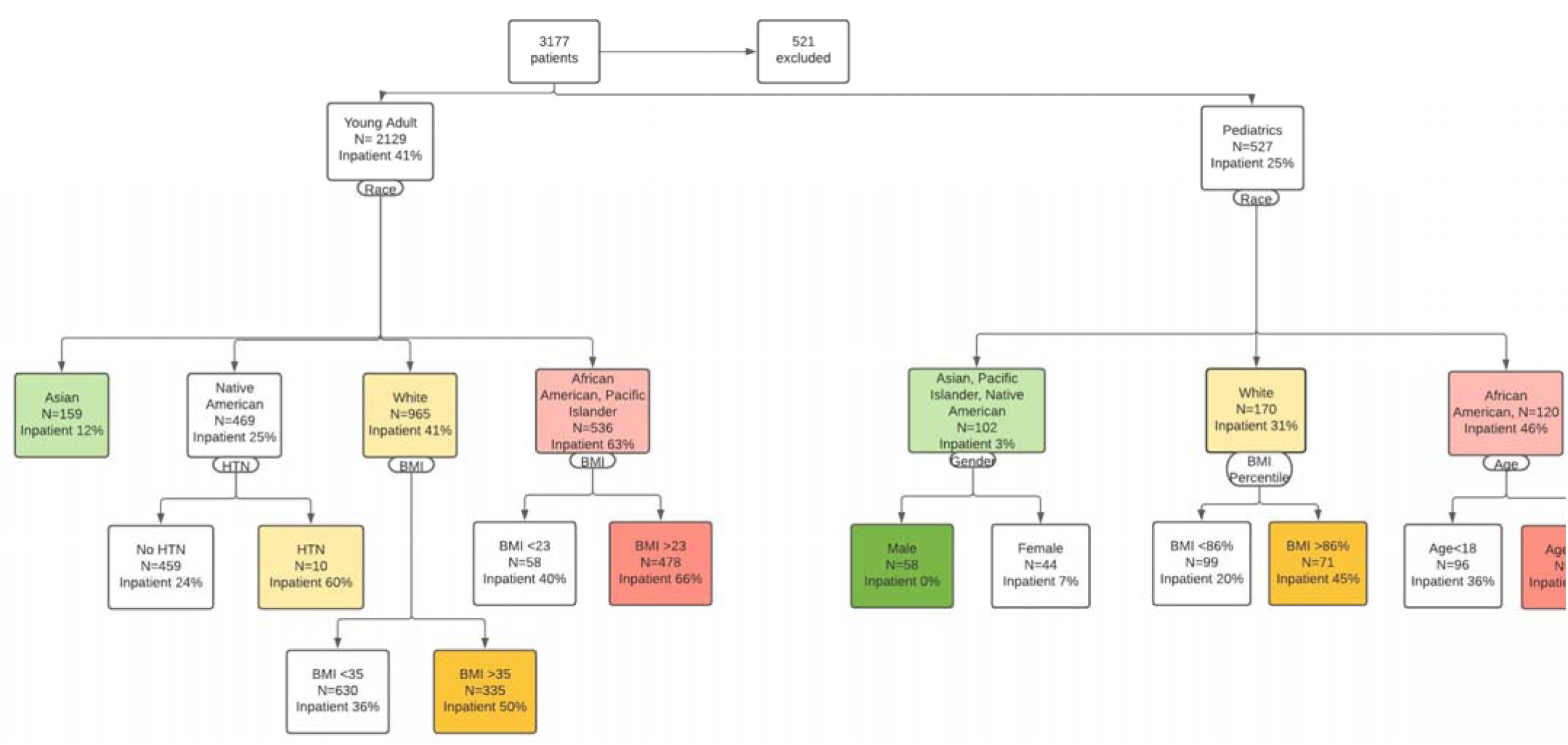
Recursive Partitioning Results for Young Adult and Pediatric Population.

For the lowest risk group, the Asians, neither BMI nor age seems to have any correlation with inpatient status.

The recursive partitioning model predicted outpatient management 99% of the time.

### Young adult population analysis

Analyzing the young adult population, we found a significant difference between BMI categories and admission status (**Table2**). Our results showed that young adults with overweight or obese BMIs had an increased likelihood of admission when compared to those with a healthy BMI (2.47, CI 1.98-3.08). No significant ventilation intervention was found between BMI categories, however out of the eight patients requiring ventilation, 2 were obese and 6 were overweight. Obesity was also associated with increased co-morbidity, 26% compared to 15.5% and 14% of overweight and healthy weight respectively. Four deaths recorded in this population were all in the obese BMI category, however the analysis didn’t show any significant relationship between BMI category and mortality.

When we looked at race, weight status and admission, we found a significant difference. The White inpatient population is shown in **Table 3** and the African American inpatient population is shown in **Table 4**. In both populations, significantly more obese people required admission. Overall, 64% of African American adults with COVID-19 were admitted compared to 39% of white adults with COVID-19.

**Table 3:**
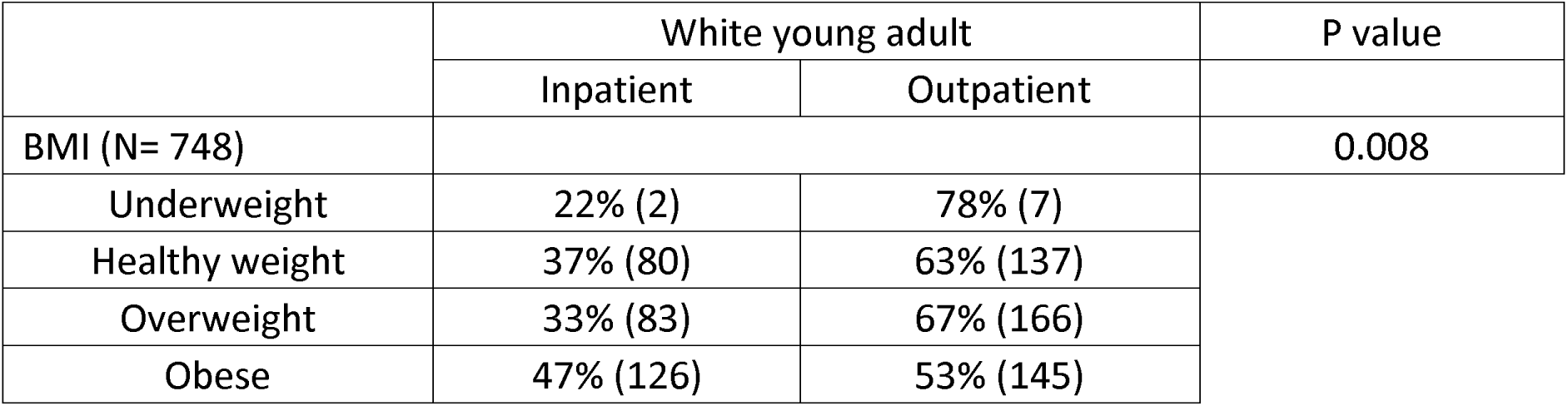
The admission distribution of each weight class of white young adult.

|  | White young adult |  | P value |
| --- | --- | --- | --- |
|  | Inpatient | Outpatient |  |
| BMI (N= 748) |  |  | 0.008 |
| Underweight | 22% (2) | 78% (7) |  |
| Healthy weight | 37% (80) | 63% (137) |  |
| Overweight | 33% (83) | 67% (166) |  |
| Obese | 47% (126) | 53% (145) |  |

**Table 4:** The admission distribution of each weight class of African American young adult.

|  | African American young adult |  | P value |
| --- | --- | --- | --- |
|  | Inpatient | Outpatient |  |
| BMI (N=431) |  |  | 0.014 |
| Underweight | 50% (2) | 50% (2) |  |
| Healthy weight | 51% (39) | 49% (38) |  |
| Overweight | 59% (61) | 41% (42) |  |
| Obese | 70% (172) | 30% (75) |  |

When analyzing using the recursive partitioning (**Figure1**), the greatest predictor for admission status was race. Like the pediatric cohort, a greater majority of African American adult were admitted. The next factor was BMI, African American with a BMI greater than 23 had a higher inpatient predictor while the White population had a greater inpatient predictor with a BMI greater than 35.

This recursive partitioning model of young adult successfully predicted outpatient stratification 87% of the time, which is slightly lower when compared to the pediatric cohort which was predictive at 99% of the time.

## Discussion

This study examines demographic characteristics and clinical course for 2131 young adult and 527 pediatric patients with COVID-19 from a population of patients from Western New York. The regional database used allowed for the analysis of a large population size with data from multiple institutions.

By applying the standard WHO definition for obesity to a large sample size in a younger patient population with fewer comorbidities, we hoped to use a reproducible standard definition in a population with a lower inherent incidence of potential confounding variables. The use of the standard WHO definition was also important in that some prior studies used different BMIs for their definition of obesity.^8,10^,

Furthermore, our population was specifically selected for their low incidence of comorbidities associated with age and obesity, which could potentially confound analysis. These variables include diabetes, hypertension, and coronary artery disease, variables which have been shown in prior studies to be independently associated with more severe clinical courses for patients with COVID 19.^3,4,5,6,7^ By choosing a population with inherently few comorbidities, we hoped to address whether obesity could be considered an independent epidemiological risk factor for admission in patients with COVID-19.

The use of recursive partitioning allowed for the analysis of patients with missing data while providing a model that examines race and ethnicity while adjusting for medical complexity. We examined the need for inpatient versus outpatient management as a surrogate for severity of clinical course. While linear regression identified race, BMI, smoking status, and hypertension as statistically significant risk factors predicating admission, we used recursive regression to help quantify the relative importance of these risk factors and develop models that were accurate at predicting patients managed as outpatients.

Our results showed that young adults with overweight or obese BMIs had an increased likelihood of admission when compared to those with a healthy weight. In this population, our results suggest a further interplay between race and obesity, as lower BMIs were associated with increased risk of admission for African Americans than for Whites.

Interestingly, obesity was not an independent risk factor for admission in our pediatric population, despite being a risk factor for disease severity in other studies, including a recent study in which obesity was a noted comorbidity in a population of 48 pediatric patients admitted to the Pediatric Intensive Care Unit.^17^

As the COVID 19 pandemic as progressed, racial disparities in the population impact and clinical course of SARS CoV2 /COVID 19 have grown more apparent. ^11^ ^12, 14^ Communities that are primarily black have been shown to have a much higher rate of COVID-19 cases than communities that are primarily white.^15^ Our data was collected primarily from residents of the 8 counties of Western New York. Of these, Erie County is the most populous of these counties, and also has the lowest percentage of white residents. According to most recent Erie County census data, Erie County is 80% White, 14% Black, 4% Asian, and 6% Latino or Hispanic. Despite this racial distribution, 25% of the young adult and 23% of pediatric population in our dataset identified as Black/African American. This has been noted in recent literature, and our results provide further support for this known disparity^15^. Importantly, these populations also have higher baseline rates of hypertension, diabetes, coronary artery disease, and obesity; all of which are established risk factors for a more severe clinical course for patients with COVID-19.^6,7,13^

Our study was designed to examine a population with fewer comorbidities and therefore minimize potential confounding variables associated with obesity. By doing so, we inadvertently created a population with limited-race associated comorbidities. Through use of recursive partitioning, we then found that, for our population, race, independent of known associated comorbidities, was the strongest predictor in determining inpatient versus outpatient management of patients with COVID-19. Putative explanations for this isolated racial disparity include the fact that individuals of African American race and Hispanic ethnicity are more likely to work essential jobs and have more densely populated living conditions.^11^ This data also reflects an increasingly recognized understanding that this pandemic has disproportionately affected people of color in the United States. This in turn should provide further impetus for directing initiatives focused on recovery to these disproportionately affected groups.

## Limitations

This study was affected by several limitations. First, was the fact that, as a retrospective cohort study using a regional database, individual patient data was often incomplete. Of the 3177 patients with COVID-19, 521 (16%) were excluded for not having BMI data. For patients with BMI data available, recursive partitioning was done as an attempt to account for as many patients as possible with only partial data.

This study is also limited in that there was a low incidence of mortality and of patients requiring ventilators, and so provides a limited analysis of the full spectrum of COVID-19 disease severity.

However, arguably the most glaring limitation of this study is that the number of individuals who identify as Hispanic or LatinX is unknown. Because the United States Federal Government currently defines Hispanic as an ethnicity, and not one of the current five minimum racial categories, this information was not collected in our regional database, although it is available in local census results. In a recent Pew Research Center Survey, over 56% of Hispanic adults identified being Hispanic as part of both their racial and ethnic background.^18^ This serves as a stark reflection of how current categories are inadequate at capturing a multifaceted question, and current tools are neither comprehensive or standardized. Where data for Hispanic/LatinX individuals has been available, it has shown that this population, like African Americans, has been disproportionately impacted by COVID 19.^11^ From a larger perspective, this limitation is in itself hugely significant as it provides a clear example of a systems level obstacle to obtaining basic data needed to inform and drive potential interventions for racial/ethnic minorities shortchanged by the current social, economic, and medical system. For our specific population, it is unknown if individuals who self-identify as Latino/LatinX would have selected ‘Other’ or ‘White’ in the data collected. As a result, it is a potential major limiting factor in our results.

## Conclusion

Our study suggests that race is the most important determinant in predicting the need for inpatient versus outpatient management of young adults and children with COVID-19. Obesity, as determined by the WHO classification system, independent of associated comorbidities, was less significant in our young adult, and insignificant in out pediatric population. Future studies should focus on examining more closely the impact COVID-19 has had on Hispanic/LatinX populations, and the significance of race in older patient populations. When determining the distribution of resources dedicated to prevention, management, and medical and economic recovery efforts, the difference in impact between races should be considered.

## Data Availability

All data produced in the present study are available upon reasonable request to the authors

## Notes

### Competing Interest Statement

The authors have declared no competing interest.

### Funding Statement

This study did not receive any funding

### Author Declarations

University at Buffalo Institutional Review Board (UBIRB) gave ethical approval for this work

